# How to remove the testing bias in CoV-2 statistics

**DOI:** 10.1101/2020.10.14.20212431

**Authors:** Klaus Wälde

## Abstract

**BACKGROUND:** Public health measures and private behaviour are based on reported numbers of SARS-CoV-2 infections. Some argue that testing influences the confirmed number of infections.

**OBJECTIVES/METHODS:** Do time series on *reported* infections and the number of tests allow one to draw conclusions about *actual* infection numbers? A SIR model is presented where the true numbers of susceptible, infectious and removed individuals are unobserved. Testing is also modelled.

**RESULTS:** Official confirmed infection numbers are likely to be biased and cannot be compared over time. The bias occurs because of different reasons for testing (e.g. by symptoms, representative or testing travellers). The paper illustrates the bias and works out the effect of the number of tests on the number of reported cases. The paper also shows that the positive rate (the ratio of positive tests to the total number of tests) is uninformative in the presence of non-representative testing.

**CONCLUSIONS:** A severity index for epidemics is proposed that is comparable over time. This index is based on Covid-19 cases and can be obtained if the reason for testing is known.

## 1 Introduction

### Background

Statistics have gained a lot in reputation during the Covid-19 pandemic. Almost everybody on this globe follows numbers and studies “the curve” on recorded cases, on daily increases or on incidences of CoV-2 infections.

### The open question

What do these numbers mean? What does it mean that we talk about “a second wave”? Intuitive interpretations of “the curve” suggest that the higher the number of new infections, say in a country, the more severe the epidemic is in this country. Is this interpretation correct? When the number of infections increases, decision makers start to discuss additional or tougher public health measures. Is this policy approach appropriate?

### Our message

Reported numbers of CoV-2 infections are probably not comparable over time. When public health authorities report *x* new cases on some day in October 2020, these *x* new cases do not have the same meaning as *x* new cases in April, May or June 2020. The bias results from different testing rules that are applied simultaneously. Private and public decision making should not be based on time series of CoV-2-infections as the latter do not provide information about the true epidemic dynamics in a country. If the reason for testing was known, a unbiased measure of the severity of an epidemic could be computed easily.

### Our framework

We present a theoretical framework that allows one to understand the link between testing and the number of reported infections. We extend the classic SIR model (Kermack and McKendrick, 1927, Hethcote, 2000) to allow for asymptomatic cases and for testing.^2^ Our fundamental assumption states that the true numbers of susceptible, infectious and removed individuals are not observed.

### Results

The reason for the intertemporal bias consists in relative changes of test regimes. If a society always employed only one rule when tests are taken, e.g. “test for SARS-CoV-2 in the presence of a certain set of symptoms”, then infection numbers would be comparable over time. If tests are undertaken simultaneously, e.g. “test in the presence of symptoms” but also “test travellers without symptoms”, and the relative frequency of tests changes, a comparison of the number of reported infections over time bears no meaning. The paper illustrates the bias by a “second wave” in reported cases which - by true epidemiological dynamics - is not a second wave.

Understanding this bias also provides an answer to one of the most frequently asked question when it comes to understanding reported infection numbers: What is the role of testing? Do we observe a lot of reported infections only because we test a lot? Should we believe claims such as “if we test half as much, we have half as many cases”. This paper will provide a precise answer to what extent the reported number of infections is determined by the number of tests in a causal sense. The answer in a nutshell: If tests are undertaken because of symptoms, there is no causal effect from the number of tests on the number of reported infections. If tests are undertaken for other reasons (travellers, representative testing), the number of reported infections go up simply because there is more testing.^3^

We show that time series on the number of tests and time series on reported infections do not allow one to obtain information about the true state of an epidemic. We also study the positive rate as the ratio of the number of positive tests to the total number of tests. The positive rate is informative if we undertook representative testing only. The positive rate is not informative about true epidemiological dynamics when there are several reasons for testing.

Understanding the biases also allows us to understand how to correct for it. The paper presents a severity index for an epidemic that is unbiased. One can obtain this index in two ways: Record the reason why a test was undertaken or count only the Covid-19 cases. Such an index should be used when thinking about relaxing or reimposing public health measures. Testing is important for detecting infectious individuals, counting Covid-19 cases is important for private and public decision making.

### Structure of paper

The next section presents the model. Section 3 shows biased and unbiased measures of the true but unobserved dynamics of an epidemic. It also studies the (lack of) informational content of time series on reported infections and time series on the number of tests, and the properties of the positive rate. It finally presents an unbiased severity index. The conclusion summarizes.

## 2 The model

The basic assumption of our extension of the susceptible-infectious-removed (SIR) model consists of the belief that true infections dynamics are not observable. Simultaneous testing of an entire population or weekly representative testing is not feasible - at least given current technological, administrative and political constraints. This section therefore first describes the true but unobserved infection dynamics, then introduces tests into this framework and finally computes the number of reported infections within this framework.

### 2.1 True but unobserved infection dynamics

#### The classic SIR model

We study a population of fixed size P. Individuals can be in three states as in a standard SIR model. The number of individuals that are susceptible to infection is denoted by 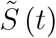. This number is unobservable to the public and to health authorities. The numbers of infectious and removed (i.e. recovered or deceased) individuals are denoted by *Ĩ*(*t*) and 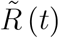, respectively. We assume that individuals are immune and non-infectious after being removed.

Let the (expected) number of individuals in the state of being susceptible at a point in time *t* be denoted by 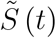.^4^ The number of susceptible individuals falls according to

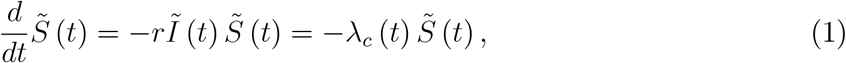

where *r* is a constant and *λ*_*c*_ (*t*) ≡ *rĨ*(*t*) can be called the individual infection rate. It captures the idea that the risk of becoming infected is the greater, the higher the number of infectious individuals.^5^ Merging individual recovery rate and death into one constant *ρ*, the number of infectious individuals changes according to

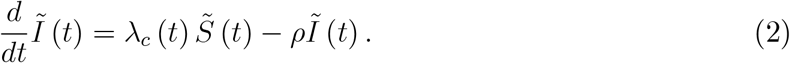

Finally, as a residual, the number of removed individuals rises over time according to 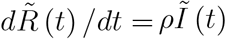. We illustrate the dynamics in the following figure, employed also later on.

#### Modelling symptomatic and asymptomatic cases

We can easily integrate asymptomatic cases into this framework. We split the true number of infectious individuals described in (2) into symptomatic and asymptomatic cases,

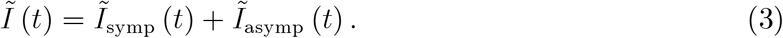

This allows us to capture the infection process in (2) by two distinct differential equations. When

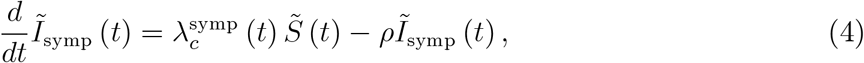

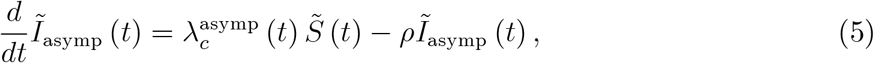

hold, (2) holds as well. Individual infection rates are now defined as

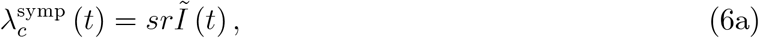

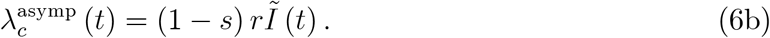

The epidemiological idea behind these equations is simple. The rate with which one individual becomes infected is the same for everybody and given by *rĨ*(*t*). The higher the number of infectious individuals in society, *Ĩ*(*t*), the higher the rate with which one individual gets infected. It then depends on various, at this point partially unknown, physiological conditions of the infected individual whether they develop symptoms or not. We denote the share of individuals that develop symptoms by s. We assume this share is constant.^6^

#### Epidemiological dynamics

This completes the description of the model. Let us now describe how we can understand (unobserved) epidemiological dynamics. We start with some initial condition for 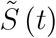. A good candidate would by 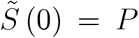, i.e. the entire population of size *P* is susceptible to being infected and become infectious. Initially, there are very few infectious individuals, say, there are two, *Ĩ*_*symp*_ (*t*) = *Ĩ*_*asymp*_ (*t*) = 1. Given infection rates (6a) and (6b) and parameters, the number of infectious symptomatic and asymptomatic cases evolves according to (4) and (5). Infectious individuals are removed from being infectious at a rate *ρ*.^7^ The number of susceptible individuals follows (1).

The epidemic is over with herd immunity, i.e. 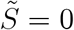 at some point (far in the future) or when recovery is sufficiently fast relative to inflows such that *Ĩ*= 0.^8^ The epidemic is heading towards an end when 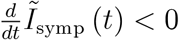 and 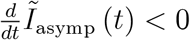, i.e. the number of infectious individuals falls.^9^ We abstract from public health measures and their effects (as studied e.g. by Dehning et al., 2020 or Donsimoni et al., 2020a, b,). If we wanted to include them, we could allow public health measures to affect r in the individual infection rate in (2).^10^

### 2.2 Modelling tests for SARS-CoV-2

To understand the effects of tests, we now introduce testing into our SIR model. The following figure displays all unobserved quantities in the model by dashed lines. The red circles represent the standard SIR model illustrated in figure 1.

**Figure 1.**
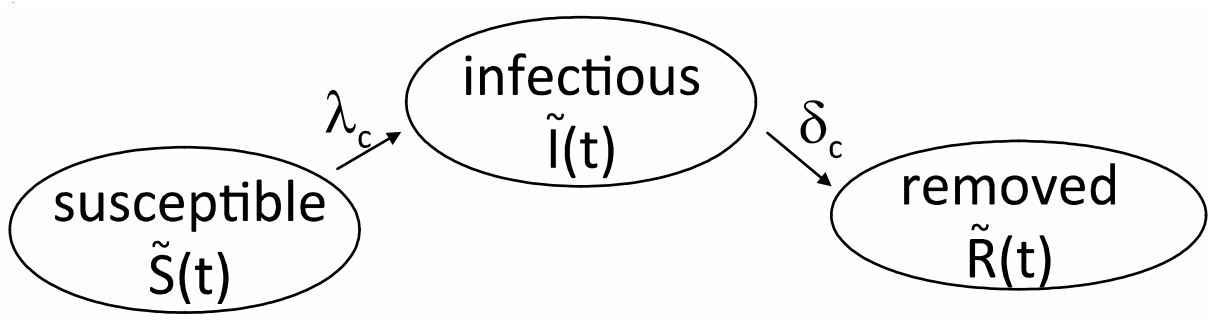
The true infection dynamics (a simple SIR model)

Testing can take place for a variety of reasons described in test strategies adopted by various countries. The reasons for tests we take into account at this point is testing due to the presence of typical symptoms, representative testing and testing travellers. While testing by symptoms and representative testing is well-defined, testing travellers is really only an example for a larger type of test. This example covers all tests that are applied to a group defined by certain characteristics which, however, are not representative of the population as a whole. Other examples of this non-representative testing include testing of soccer players, testing in retirement homes or their visitors, testing in hot spots or testing contact persons of infected individuals.

#### Testing by symptoms

Individuals can catch many diseases (or maybe better sets of symptoms) indexed by *i* = 1…*n*. For simplicity, the above figure displays only two diseases (1 and 2) and Covid-19. The number of individuals that have a disease *i* and go to a doctor on day *t* is *D*_*i*_ (*t*). An individuals becomes sick with an arrival rate *λ*_*i*_ and recovers from this specific sickness *i* with a rate of *ρ*_*i*_. For clarity, we add a symptomatic SARS-CoV-2 infection to this list of diseases. The individual is infected and develops symptoms with rate 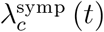, which we know from (6a), and is removed with rate *ρ*_*c*_. The number of symptomatic SARS-COV-2 individuals is *Ĩ*_*symp*_ (*t*) from (4).

There is a certain probability *p*_*i*_ that a doctor performs a test, given a set of symptoms *i*. This probability reflects the subjective evaluation of the general practitioner (GP) whether certain symptoms are likely to be related to SARS-CoV-2. The probability to get tested with symptomatic SARS-CoV-2 infection (which the GP of course does cannot diagnose without a test) is denoted by p_c_. Hence, the (average or expected) number of tests that are performed at time t due to consulting a doctor is given by

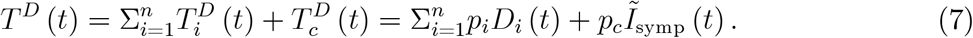

The second equality replaces the number of tests by the number of sick individuals per disease times the probability that this individuals is tested.^11^

Note that, apart from population size P, the number of tests taken because of the presence of symptoms, *T*^*D*^ (*t*), is the first variable that is observed. If health authorities collected information why a test was performed (set of symptoms that can be observed by a GP), we would observe 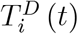 and 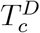. If not, we observe *T*^*D*^ (*t*) only.

#### Testing travellers or for scientific reasons

Tests can be performed for a variety of reasons. One consists in testing travellers, another consists in tests for scientific reasons and so on. Theses tests are not related to symptoms. Taking the example of representative tests, the tests are applied to the population as a whole. The number of tests is chosen by public authorities, scientists, available funds, capacity considerations and other. In any case, it is independent of infection-characteristics of the population. Concerning representative testing, we denote the number of tests of this type undertaken at *t* by *T*^*R*^ (*t*). When it comes to travelers, we denote the number of tests by *T*^*T*^ (*t*).

Summarizing, the total number of tests being undertaken in our model is given by the sum of tests due to symptoms, *T*^*D*^ (*t*), representative tests *T*^*R*^ (*t*) and testing travellers, *T*^*T*^ (*t*),

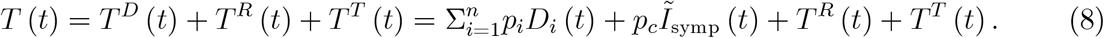

The second equality employs the number of tests by symptoms from (7). The equation thereby reemphasizes the endogeneity of the number of tests by symptoms, *T*^*D*^ (*t*) is determined by the number of symptoms occurring in a country or region, and the exogeneity of other reasons for testing, *T*^*R*^ (*t*) and *T*^*T*^ (*t*). The latter are not determined by symptoms.

### 2.3 The number of reported infections

The number of reported infections at time *t* is given by the sum of reported infections split by the reasons for testing introduced above,

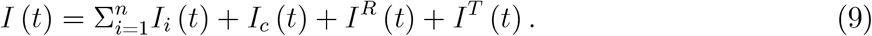

#### Testing by symptoms

As we are perfectly informed in our theoretical world about the (expected) number of CoV-2 infections and other diseases, we know that the number of positive CoV-2 tests is zero for all diseases,^12^

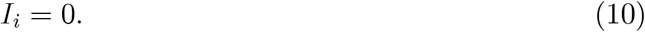

Individuals have Covid-19 related symptoms because they caught a cold, they have the flu or other. The probability that a CoV-2 infected individual has a positive test is set equal to one (ignoring false negative tests). The number of positive tests for individuals that are infected with CoV-2 is therefore identical to the number of tests,

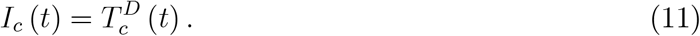

#### Testing for other reasons

The probability that a representative test is positive is denoted by *p*^*R*^ (*t*). This probability is a function of the true underlying and unobserved infection dynamics. If the sample chosen is truly representative, then the probability for a positive test is given by

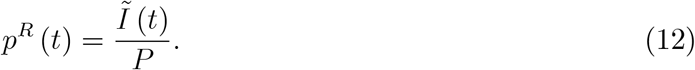

Hence, representative tests make the true number *Ĩ*(*t*) of infectious individuals visible for the moment at which the tests are undertaken. This true number includes symptomatic and asymptomatic cases as in (3).

The probability that a test of travellers is positive depends on a multitude of determinants among which region traveled to and behaviour of the traveller. We denote the probability that such a test is positive by *p*^*T*^ (*t*). We consider this probability to be exogenous to our analysis.

#### Total reported infections

A first step towards the total number of reported infections starts from (9) and takes (10) and (11) into account,

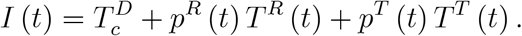

This is also the expression displayed in figure 2 between ‘CoV-2 tests’ and ‘confirmed infections’. Reported infections come from testing CoV-2 individuals with symptoms, from representative testing and from other sources such as travellers. Employing 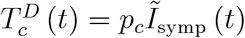 from (7) and *p*^*R*^ (*t*) from (12), the number of reported infections can be written as

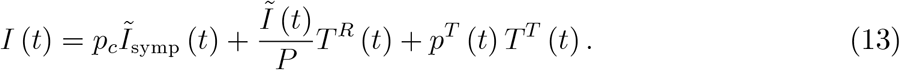

**Figure 2.**
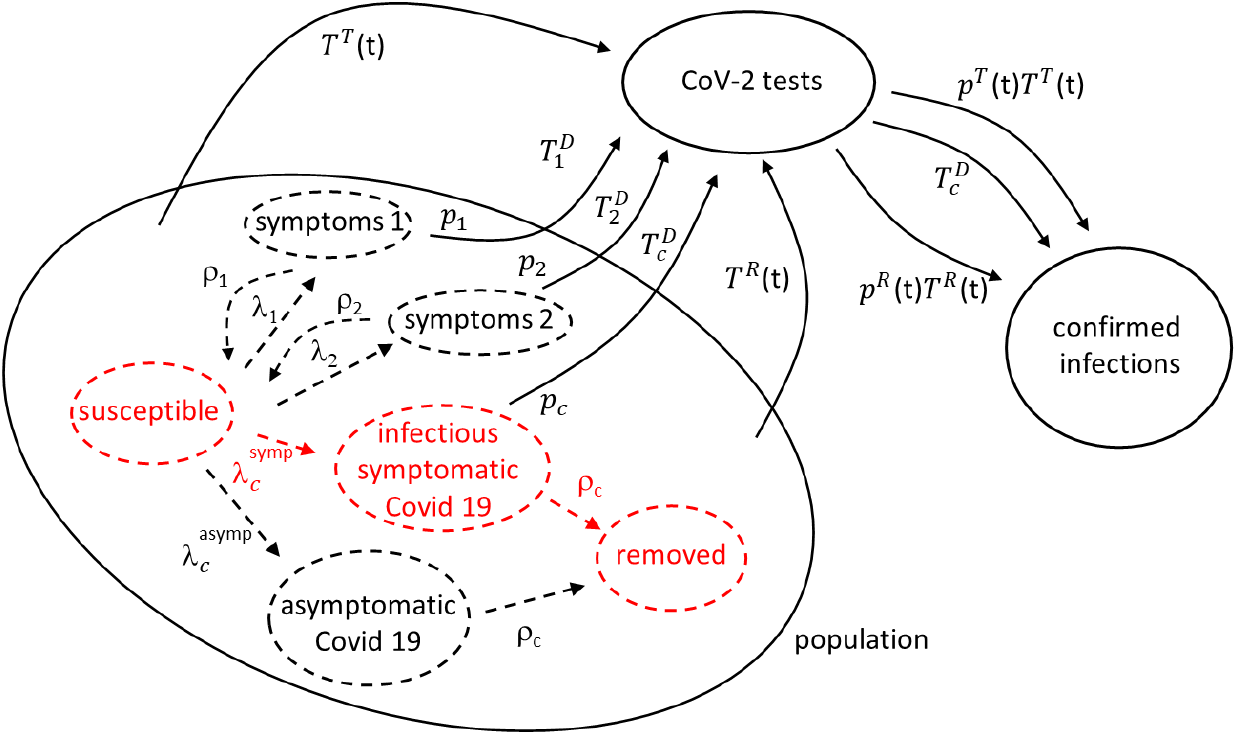
The SIR model with testing

## 3 Unbiased and biased reporting

Before we compute the bias in the number of infections, let us define what we understand by an unbiased time series of numbers of infections. A time series is unbiased if it is proportional to the true (but unobserved) number of infections. An example of a biased time series is then simple: If the number of tests increases the number of reported infections, then the number of reported infections is not informative about the true number of infections.

### 3.1 Unbiased reporting

Our central equation is (13). The reported number of infections would be unbiased if only tests by symptoms were undertaken. Reported infections (with *T*^*R*^ (*t*) = *T*^*T*^ (*t*) = 0) would by (13) amount to

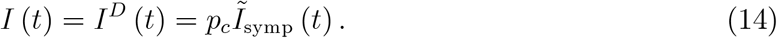

When the reported number of infections *I*(*t*) goes up, one would be certain that the unobserved number of symptomatic CoV-2 infections *Ĩ*_*symp*_ (*t*) would go up as well. The more infections are reported, the more severe the epidemic is.

This equation also shows under which circumstances the number of tests does *not* have a causal effect on the number of reported infections. If tests are undertaken according to a rule that makes testing dependent on something else (e.g. the presence of symptoms), the number of tests itself is determined by the number of symptoms. Hence, while the number of tests and reported infections are correlated, the causal underlying factor is the number of patients visiting a physician with CoV-2 related symptoms.

A second example of unbiased testing is (exclusive) representative testing. When only representative testing is undertaken, the number of reported infections (with 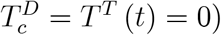 from (13) amounts to

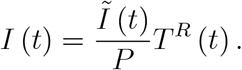

Here, the number of reported infections, *I*(*t*), does rise in the number of tests, *T*^*R*^ (*t*). The more we test, the higher the number of cases. Yet, representative testing is (of course) the gold standard of testing. The ratio of positive cases to the number of tests yields the share of infections in the population,^13^

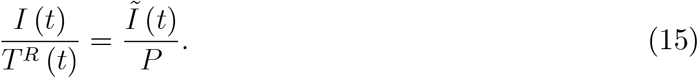

This share is driven by *Ĩ* (*t*) which shows that (i) representative testing provides a snapshot at this point in time *t* of the current epidemic dynamics and that (ii) representative testing provides a measure of overall infections, i.e. symptomatic and asymptomatic ones.

We have seen two examples of unbiased reporting, one for symptomatic infections, one for all infections. They show that the question whether the number of reported cases rises in the number of tests is not as important as the question whether the type of testing provides useful information. In the first example, the claim that more tests increase the number of reported infections is meaningless as the number of tests is not chosen. In the second example, the number of positive cases rises in the number of tests but the ratio of these two quantities is highly informative.

### 3.2 Biased reporting

#### Illustrating a bias

Now imagine several types of testing are undertaken simultaneously. The number of reported infections at *t* is then given by the full expression in (13). Consider first the case of symptomatic and representative testing. The number of reported cases (with *T*^*T*^ (*t*) = 0) is then

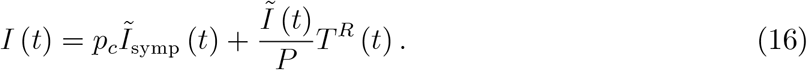

Imagine someone (the government, researchers, other) decide to undertake more representative testing, i.e. *T*^*R*^ (*t*) goes up. This means that *I*(*t*) increases even though there is no change in the true number *Ĩ*_*symp*_ (*t*) of symptomatic cases. There is also no change in the true number *Ĩ*(*t*) of symptomatic and asymptomatic cases. Whoever perceives the reported number *I*(*t*) is led to believe that something fundamental has changed within the epidemiological dynamics. But this is of course not true. The reported number goes up simply because more tests were undertaken.^14^

Can we gain some information out of this expression if we divide it by the number of tests *T*^*R*^ (*t*) as it had turned out to be very useful in the case of exclusive representative testing in (15)? We would obtain

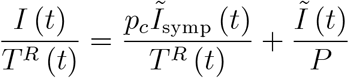

which does contain the informative infection share *Ĩ*(*t*) /P as the second term on the right hand side. But the first term does not have a meaningful interpretation and neither does the entire term.

#### A numerical example of a bias

Let us illustrate the potential bias by looking at the third type of testing considered here - testing travellers. The number of reported infections according to (14) in the case of testing by symptoms and testing travellers reads

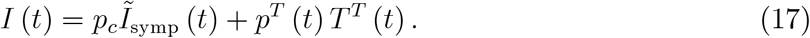

We assume that no testing of travellers took place at the beginning of the pandemic. At some later point (as of *t* = 60 in our figure below), the number of tests per day, *T*(*t*), increases linearly in time.

To make this example as close to public and common displays of infection dynamics, let us look at “the curve” represented in figure 3 by numbers of infected individuals taking recovery into account.^15^

**Figure 3.**
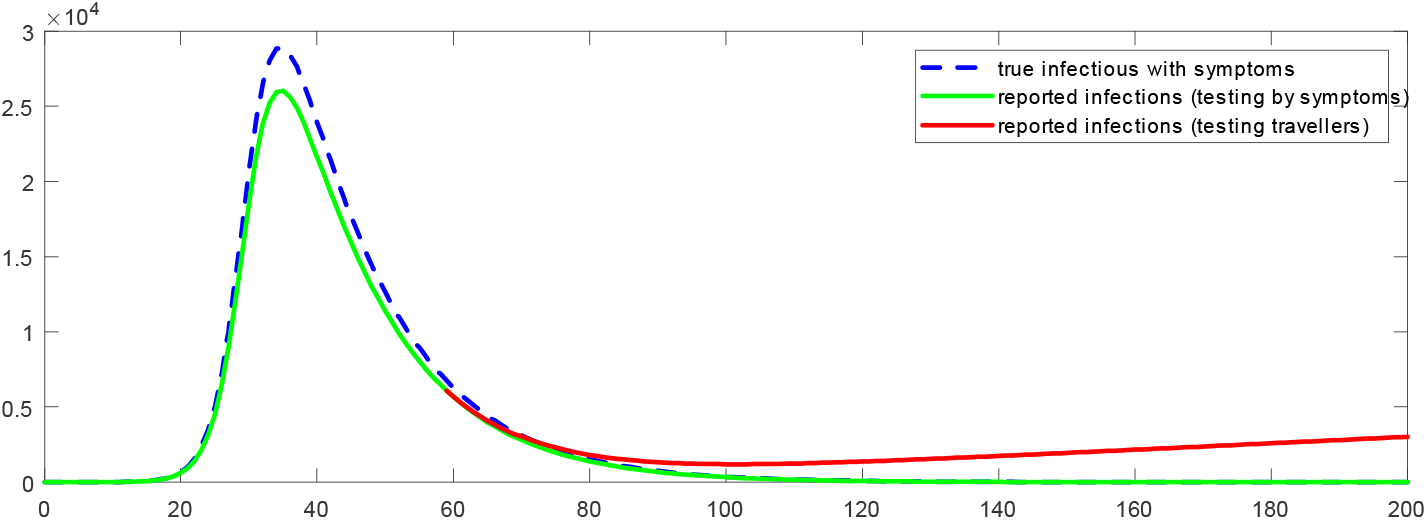
True epidemiological dynamics (blue) correct reporting (green) and an example of a bias (red) of the reported number of infections

Looking at figure 3 we first focus on the blue dashed curve for *Ĩ*_*symp*_ (*t*), the true number of symptomatic SARS-CoV-2 infections. We chose parameters such that the epidemic is coming to a halt after around 120 units of time (plotted on the horizontal axis). The green curve plots the number of reported infections *I*^*D*^ (*t*) from (14) where testing takes place only in the presence of symptoms. Finally, the red curve is an example of a bias in the reported number of infections. It occurs as positive tests from testing travellers are added to tests by symptoms as in (17).

We see that this example displays what looks like a “second wave”: reported numbers of infections go up again as of *t* = 100. By construction, however, this second wave is caused by misinterpretation of the reported number of infections. Let us stress that we do not claim that the second wave is a statistical artefact due to testing strategies. It could be a statistical artefact, however. The conclusion shows how to obtain a severity index for an epidemic that is not prone to causing artificial results and which data is needed to compute such an index.

### 3.3 Two non-applications

#### A non-application to Germany

Consider the case of Germany. Figure 4 shows the number of tests per week and the number of reported infections. When we look at the time series for all tests in this figure, it corresponds to *T* (*t*) from (8). When we consider the reported number of infections per week in Germany, it looks as displayed in the right panel of the above figure. This time series corresponds to *I*(*t*) from (13). Can we conclude anything from these two time series about the true dynamics of the epidemic, i.e. can we draw conclusions about *Ĩ*_*symp*_ (*t*) or *Ĩ*(*t*)?^16^

**Figure 4.**
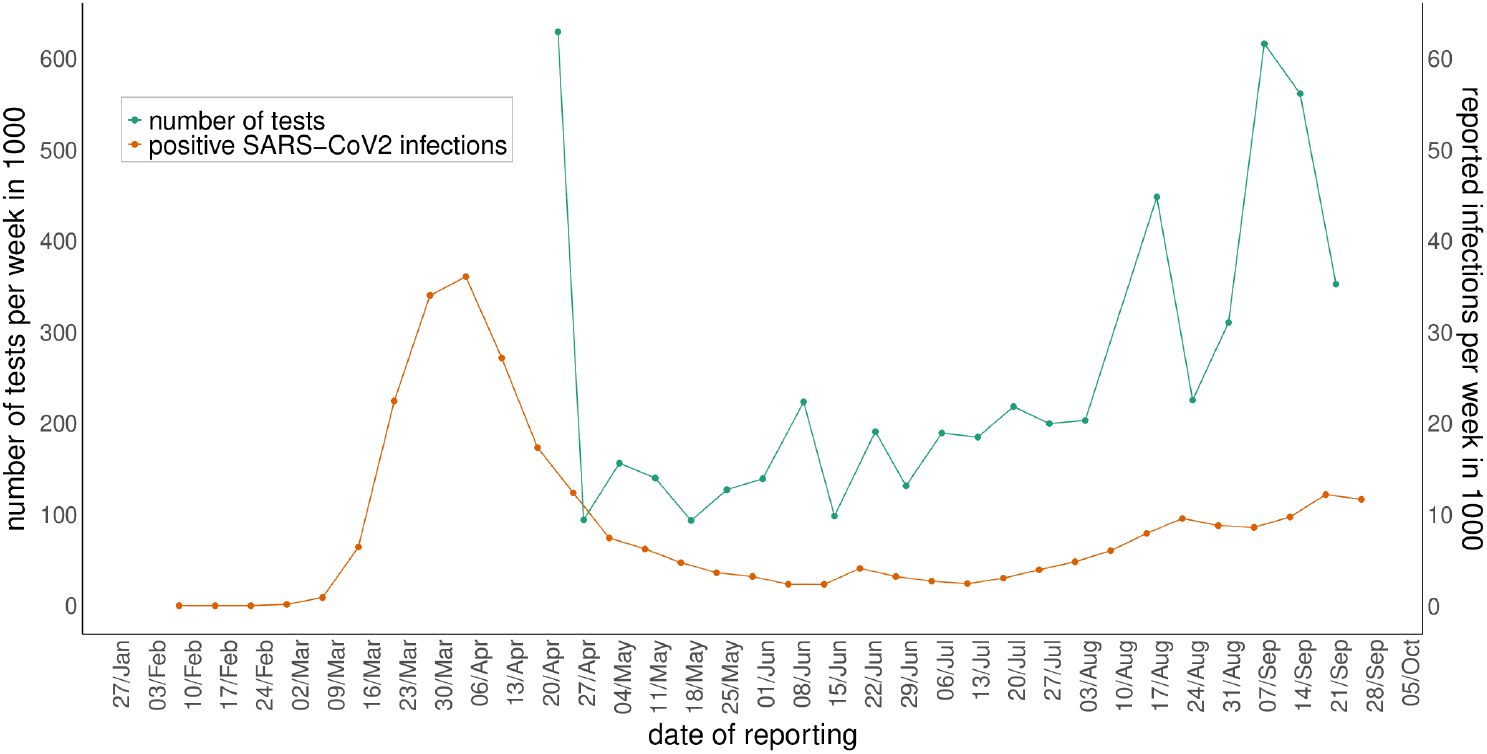
The number of weekly tests and infections in Germany

Technically speaking, we have two equations, (8) and (13), reproduced here for convenience,

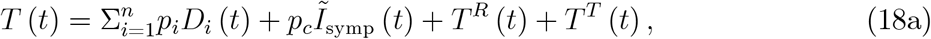

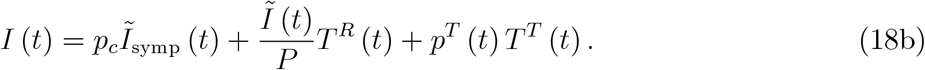

about which the public has access to two variables, *I*(*t*) and *T* (*t*). It seems obvious that - unless we want to make a lot of untested assumptions - official statistics do not allow to draw any conclusion about the severity of the epidemic. The right-hand side contains at least three unknowns (e.g. tests classified by reason of testing, *T*^*D*^ (*t*), *T*^*R*^ (*t*), *T*^*T*^ (*t*)) and two equations with three unknowns usually do not have a solution. Hence, from currently available data, true epidemic dynamics, *Ĩ*_*symp*_ (*t*) or *Ĩ*(*t*), cannot be understood.^17^

#### The positive rate

The positive rate is the ratio of confirmed infections to number of tests, *s*^*pos*^ (*t*) ≡ *I*(*t*) /*T*(*t*). This statistic is often discussed in the media and elsewhere (see e.g. Our World in Data, 2020). In our model, (13) and (8) imply

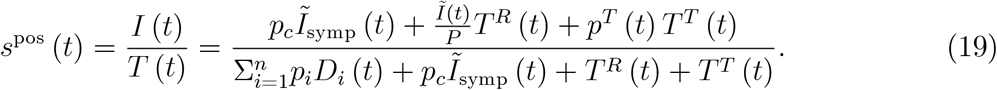

What does this positive rate tell us? Some argue that a rising positive rate is a sign of the epidemic ‘getting worse’. If we understand the latter by a rise in the number of unobserved infections, *Ĩ*(*t*), or the number of infections with symptoms, *Ĩ*_*symp*_ (*t*), this statement is true if we undertake representative testing only, 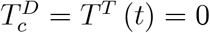 as in (15). In this case,

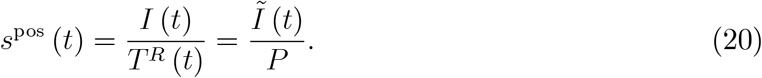

When the observed positive rate *s*^*pos*^ (*t*) rises, this clearly indicates that the number of unobserved infections *Ĩ*(*t*) is higher. And so would be *Ĩ*_*symp*_ (*t*), whether individuals with symptoms go to a doctor or not, given the constant share s of symptomatic cases in (6a).^18^

Does this conclusion hold more generally, i.e. for the full expression (19) when tests are undertaken for many reasons? Let us assume we only undertake tests due to symptoms and due to travelling, *T*^*R*^ (*t*) = 0.^19^ Then the positive rate (19) reads

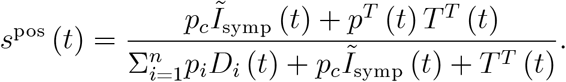

When we increase the number of tests for travellers, we find (see appendix)

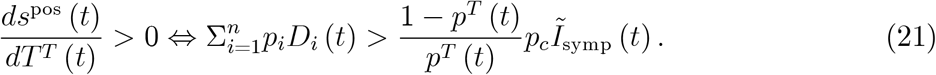

This result is easy to understand technically and has the usual structure: When we increase a summand (*T*^*T*^ (*t*) here) that appears in a fraction in numerator and denominator, the sign of the derivative depends on the other summands (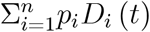 and *p*_*c*_*Ĩ*_*symp*_ (*t*) in this case). As the summand is multiplied by *p*^*T*^ (*t*) in the numerator, this probability appears in the condition as well.

In terms of epidemiological content, the derivative says that the positive ratio can rise or fall when we increase the number of tests for travellers (or related reasons mentioned below figure 2). Testing increases the positive rate if the number of tests undertaken due to symptoms that are not CoV-2 related, 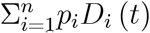, exceeds the number of tests undertaken because of symptoms related to CoV-2, *p*_*c*_*Ĩ*_*symp*_ (*t*), corrected for the probability that a traveller test is positive.

While an intuitive interpretation of this condition seems to be a challenge, the condition nevertheless conveys a clear message: It contradicts that a rising positive rate implies a ‘worse’ epidemic state. We see that when *T*^*T*^ (*t*) goes up and the positive rate goes up, this does not mean anything regarding the dynamics of *Ĩ*_*symp*_ (*t*) or *Ĩ*(*t*). The same is true, of course, when *T*^*T*^ (*t*) goes up and the positive rate goes down. The positive rate is not informative.

This finding also applies to a somewhat more precise statement of the above conjecture. Some claim that a rising positive rate in the presence of more tests does show that infections must go up. When we increase *T*^*T*^ (*t*), the number of tests goes up. When (21) holds, the positive rate goes up. However, we do not learn anything about infections with or without symptoms. Tests go up and the positive rate goes up simply because we test more.

### 3.4 An unbiased severity index for an epidemic

#### A very simple index

We now propose an index for the severity of an epidemic which is comparable over time. The model illustrated in figure 2 tells us what is needed: The index should be closely related to the number of symptoms in society. As tests that capture these symptoms are those that are undertaken because of symptoms, the index is simply *I*^*D*^ (*t*) as in (14).

An alternative would consist in representative testing. While the number of reported cases depends causally on the number of tests, the ratio of reported cases to number of tests is an unbiased estimator of the true epidemic dynamics as shown in (15). As regular representative testing, say with a weekly frequency, is not feasible, the only realistic severity index is *I*^*D*^ (*t*) from (14).

Very simply speaking: If a severity index for an epidemic is desired that is comparable over time, we should test for CoV-2 but count Covid-19 cases. This should be done at all levels starting from the GP, through hospital admissions and patients in intensive care and, finally, counting deaths associated with Covid-19.

#### Policy lessons

What do these findings mean in practice? Data which is currently available for the public (see e.g. RKI, 2020 for Germany or Our World in Data, 2020, for many other countries in the world) does break down the total numbers of tests by origin (GP, hospital and other) and region. Unfortunately, this classification does not relate to the reason for testing and the latter is required to infer the true infection dynamics.

What should be done to quantify the relevance of the bias? Local health authorities in Germany collect the names of individuals with confirmed CoV-2 infections. If additional information on symptoms, that is *already being collected* (the reporting form^20^ allows for ticks on fever, coughing and the like), was made available to the public or scientists, the bias could be computed easily.^21^ We currently know Covid-19 cases for intensive care in hospitals, but this data is not yet easily accessible (see https://www.intensivregister.de). While only a fraction of Covid-19 cases ends up in intensive care, this number might be more informative than CoV-2 infections. The number of deaths associated with Covid-19 is a further measure as would be excess mortality. While these are only partial measures of Covid-19 dynamics, Covid-19 measures (positive CoV-2 tests with symptoms, number of all Covid-19 patients in hospitals, not just intensive care) would provide a better basis for regional and local decision makers than CoV-2 infection measures.

If one day we know how strong the quantitative bias is, we could now hope that the bias is small. Then the guidance given to society by the focus on CoV-2 infections would have been correct. But even with a small bias, the focus on CoV-2 infections should stop. We know that it is not the perfect measure. It rather biases expectation building (and emotional reactions) of individuals. Hence, as soon as better Covid-19 measures are available, the CoV-2 measure should be replaced. The candidates are estimates of informative positive rates and (regional) time series on Covid-19 cases (and not CoV-2 infections). This would allow local politicians to base their decisions on intertemporally informative data, i.e. on local Covid-19 cases.

## 4 Conclusion

True epidemic dynamics are unobserved. No country, no health authority and no scientist knows the true number of CoV-2 infections with or without symptoms for a given country. This is why testing is undertaken. Testing is a means to measure true but unobserved epidemic dynamics. The *counted number* of CoV-2 infections are *not relevant* for decision making, *what matters is the true number* of CoV-2 infections. Infections and the corresponding disease spreads when the true number of infections is high, not when the counted number of infections is high.

We extend the classic SIR model to take symptomatic and asymptomatic cases into account. More importantly, we treat CoV-2 infections as unobserved in the SIR model and model testing. We allow for various reasons for testing and focus on testing due to symptoms, representative testing and testing travellers. Testing travellers is an example of non-representative and non-symptom related testing and includes the testing of sports professionals, in retirement homes or their visitors, in hot spots or contact persons of infected individuals.

We show that the presence of various reasons for testing biases the number of confirmed CoV-2 infections over time. The number of CoV-2 infections cannot be compared intertemporally. We might observe more CoV-2 infections today simply because we test more. However, the true number of infections might stay constant or even fall.

We do not claim, in any sense, that our findings have empirical relevance. We simply do not know, at least given the data that are easily accessible to the public and given the data everybody observes (number of tests and number of reported infections) and on which all public health decisions are based, what the true epidemic dynamic is. We all look at a watch and we know that it is wrong. But we do not know how much it is wrong. It may be seconds, but it can also be hours.

What are the positive lessons from this analysis? We propose an index which is unbiased over time. It is deceptively simple. Count the number of Covid-19 cases, not the number of CoV-2 infections. If we knew the number of Covid-19 cases, i.e. CoV-2 infections with severe acute respiratory symptoms (SARS), then we would know at least one part of epidemic dynamics (*Ĩ*_*symp*_ (*t*) in our model).

Let us stress that our findings are not an argument against testing. Testing is important for identifying infectious individuals. They need to stay in quarantine in order to prevent the further spread of CoV-2 infections. This helps to reduce Covid-19 cases. Testing is important - but adding up confirmed infections from all sorts of tests is misleading. As long as the public focuses on all sources of positive cases, decisions by private individuals, firms, journalists, scientists and politicians are badly informed. Emotions, decisions and behaviour are misguided. This cannot be good for public health. Decisions must be based on the number of Covid-19 cases.

## Data Availability

Descriptive statistics on the number of CoV-2 infections in Germany from RKI and number of SARS-CoV-2 tests from RKI

## Acknowledgements

We are grateful to many commentators on earlier Covid-19 work of us for insisting that the effect of the number of tests on the reported number of infections should be clarified. We would especially like to thank Steffen Altmann, Maria Balgova, Michael Berlemann, Christian Endt, Sören Enkelmann, Bodo Plachter, Matthias Reddehase, Ulf Rinne, Achim Schneider and seminar participants at IZA Bonn and JGU Mainz for comments and discussions. A video of the JGU presentation is available at https://youtu.be/1U-OID0DJR8

## Data and materials availability

All data and software code (matlab and R) are available upon request and will be made available online.

## 5 Supplementary material

This sections contains derivations for the main text.

### 5.1 The positive rate for representative testing and testing travellers

When we ignore testing by symptoms, the positive rate (19) reads

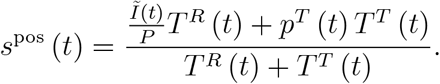

Imagine travellers were representative, then 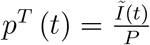 and the positive rate would read

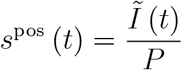

as in (20) for representative testing. Under the assumption that travellers (or visitors of retirement homes, or contact persons of a positively tested individual or visitors of public events) are representative, the positive rate would reflect the true epidemic dynamics as measured by *Ĩ* (*t*) /*P*.

### 5.2 The derivative of the positive rate in (21)

We only take tests due to symptoms and due to travelling into account, *T*^*R*^ (*t*) = 0. Then the positive rate (19) reads

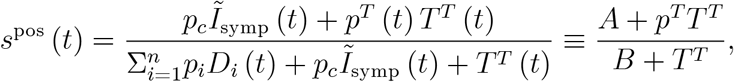

where the second equality defines A and B (for this appendix only) and suppresses time arguments to simplify notation. We compute

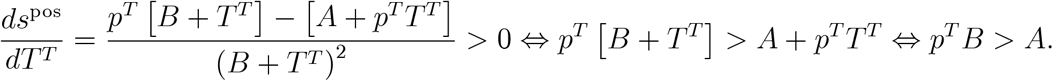

When we employ the definition of *A* and *B*, we obtain

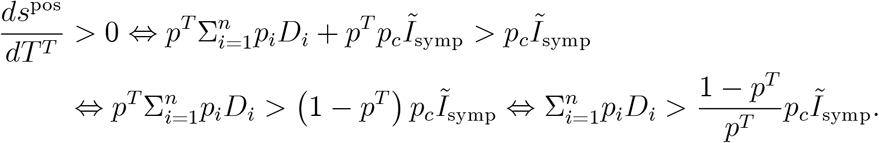

Adding time arguments gives the condition in the main text.

## Footnotes

2 Asymptomatic cases have also been taken into account e.g. by Davies et al. (2020), Ferguson et al. (2020) and Donsimoni et al. (2020). They do not analyse the biasing effect of various reasons for testing. Contributions from mathematical modelling, helping to find appropriate policy responses to pandemics, are discussed by Metcalf et al. (2020). The effect of testing strategies are not treated explicitly. We strongly share their and Robert May’s view that “mathematical models … force clarity and precision upon conjecture” (May, 2004). Testing is considered e.g. by Perkins et al. (2020). They estimate the effect of a lack of testing capacity on reported infections. They do not discuss the conceptional issue that different testing strategies bias reported infection numbers.

3 There are other sources of intertemporal biases. The speeds with which different risk groups become infected might differ, multiple testing of one person matters and general risk perceptions of medical personnel. We focus on the bias due to tests but discuss perceptions briefly below.

4 We write expected number as ordinary differential equations in SIR models could or should be understood as Kolmogorov backward equations describing means of continuous time Markov chains. See Karlin and Talyer (1998) or Ross (1996) for an introduction.

5 In the tradition of Diamand-Mortensen-Pissarides search and matching models in economics (Diamond, 1982, Mortensen, 1982, and Pissarides, 1985), this individual infection rate can be expressed capturing similar ideas as in a matching function: it should not only increase in the number of infectious individuals but also fall in the number of susceptible individuals. The latter reduces the probability that a random contact is infectious. See Donsimoni et al. (2020a) for an implementation.

6 The model neglects the effect of quarantine. If infectious individuals know about their status and therefore stay in quarantine, they should be removed from *Ĩ*(*t*) or at least get a lower weight in (6).

7 It would be straightforward to assume, e.g. *ρ*_*asymp*_ > *ρ*_*symp*_. This would capture the idea that asymptomatic cases recover faster than symptomatic cases. We ignore this extension as this distinction would not affect our main argument.

8 For analytical solutions of the classic SIR model, showing this aspect most clearly, see Harko et al. (2014) or Toda (2020).

9 This condition is related to the widely discussed reproduction number.

10 Current applications of the SIR model also badly neglect the non-exponential distribution in various states. It is well-known (e.g. Linton et al., 2020 or Lauer et al., 2020) that incubation time is (approximately) log-normally distributed. It is now also understood that the reporting delay per se and added to incubation time is also non-exponentially distributed (Mitze et al., 2020, app. A.3). The “chain trick” (Hurtado and Kirosingh, 2019) would allow to implement this numerically. Meyer-Herrmann (2018) employed a related struture but did not focus on densities of duration explicitly.

11 In a broader interpretation, one could understand *p*_*i*_ and *p*_*c*_ as the probabilities that an individual gets tested *and* that they go to the doctor. No test is ever performed if individuals with symptoms stay at home.

12 Test procedures and techniques might imply some false positive but we abstract from this at this point.

13 This ratio is an example of the ‘positive rate’. We will study it in more detail below.

14 Looking at (16) shows that a further source of bias, briefly mentioned earlier, can easily be identified. Imagine the general perception of GPs changes over time concerning Covid-19. Then a GP might be initially sceptical, i.e. *p*_*c*_ is low, then become more aware of health risks implied by CoV-2, *p*_*c*_ goes up, to then maybe during some other period become more reluctand again. If these changes in individual perceptions are not entirely idiosyncratic but driven by the overall attention in society to an epidemic, the number of reported infections would change independently of the true number of infections, *Ĩ*_*symp*_ (*t*) or *Ĩ*(*t*).

15 One could draw similar figures with new infections per day or the number of individuals ever infected. The basic argument would remain the same.

16 One might be tempted to argue that data on the positive rate in (19) should also be useful. As the positive rate is simply *I*(*t*) divided by *T* (*t*), it does not provide additional information.

17 This paper is about conceptional issues related to the finding an unbiased estimator for an unobserved time series. We ignore practical data problems. The latter include the fact that the number of tests displayed in fig. 4 is not coming from the same sample of tests that yields the number of infections in this figure. This would have to be taken into account in any application.

18 The appendix shows that the positive rate is also informative and identical to *Ĩ*(*t*) /P if travellers (or visitors of retirement homes, or contact persons of a positively tested individual or visitors of public events etc) are representative. This assumption is questionable, however.

19 Quantitatively speaking, representative testing is probably very small relative to other reasons for testing.

20 See e.g. https://www.kv-rlp.de/fileadmin/user_upload/Downloads/Mitglieder/Coronavirus/Meldepflichtige_Krankheit_Meldeformular.pdf

21 I am very grateful to Bodo Plachter for discussions of these issues and for his support.

